# Imbalanced basal ganglia connectivity is associated with motor deficits and apathy in Huntington’s disease

**DOI:** 10.1101/2021.07.01.21259448

**Authors:** Akshay Nair, Adeel Razi, Sarah Gregory, Robb R. Rutledge, Geraint Rees, Sarah J Tabrizi

**Author notes:** denotes corresponding author **Corresponding author: (1)** Dr Akshay Nair, Huntington’s Disease Centre, UCL Queen Square Institute of Neurology, University College London.

## Abstract

The gating of movement depends on activity within the cortico-striato-thalamic loops. Within these loops, emerging from the cells of the striatum, run two opponent pathways – the *direct* and *indirect* basal ganglia pathway. Both are complex and polysynaptic but the overall effect of activity within these pathways is thought to encourage and inhibit movement respectively. In Huntington’s disease (HD), the preferential early loss of striatal neurons forming the indirect pathway is thought to lead to disinhibition giving rise to the characteristic motor features of the condition. But early HD is also associated with apathy, a loss of motivation and failure to engage in goal-directed movement. We hypothesised that in HD, motor signs and apathy may be selectively correlated with indirect and direct pathway dysfunction respectively. We used spectral dynamic casual modelling of resting state fMRI data to model effective connectivity in a model of these cortico-striatal pathways. We tested both of these hypotheses *in vivo* for the first time in a large cohort of patients with prodromal HD. Using an advanced approach at the group level by combining Parametric Empirical Bayes and Bayesian Model Reduction procedure to generate large number of competing models and compare them by using Bayesian model comparison. With this automated Bayesian approach, associations between clinical measures and connectivity parameters emerge *de novo* from the data. We found very strong evidence (posterior probability > 0.99) to support both of our hypotheses. Firstly, more severe motor signs in HD were associated with altered connectivity in the indirect pathway components of our model and, by comparison, loss of goal-direct behaviour or apathy, was associated with changes in the direct pathway component. The empirical evidence we provide here is demonstrates that imbalanced basal ganglia connectivity may play an important role in the pathogenesis of some of commonest and disabling features of HD and may have important implications for therapeutics.

## Introduction

Huntington’s disease (HD) is an autosomal dominant neurodegenerative condition caused by a triplet repeat expansion in the *huntingtin* gene on chromosome 4.^1,2^ While the aetiology of HD is clear, the pathogenesis of many of the core clinical motor, cognitive and behavioural features of HD remain to be established. Although HD ultimately affects almost the entire brain, early degeneration of the striatum is canonical of this disorder both pathologically and on structural imaging.^3,4^ The striatum, however, is not a homogenous structure. As the input node to the basal ganglia it has a complex anatomy. The medium spiny neurons (MSNs) of the striatum form a wide range of compartments and pathways.^5–7^ For HD, this anatomical complexity is of relevance because the disorder does not affect all striatal MSN populations equally.^8,9^ Cortico-striatal connections, which are topographically arranged, form the input to the striatum.^10,11^ These cortical projections synapse with MSN populations that fall broadly into two key groups – those forming the *direct* and the *indirect* pathway.^12,13^ They form unique and complex polysynaptic connections with other basal ganglia structures, such as the globus pallidus, subthalamic nucleus and substantia nigra.^7^ Overall, these two pathways form opponent channels that regulate thalamic control over cortical activation.^10^ In the motor system the activity of the direct pathway encourages movement whereas the indirect pathway activity inhibits or reduces movement.^14–16^ Although all MSNs are susceptible to degeneration in HD, those of the indirect pathway appear more susceptible earlier in the disease.^9,17,18^ Based on these observations it has been hypothesised that changes in connectivity within the indirect pathway would be associated with the emergence of motor signs in HD which are characterised by erratic, noisy and disinhibited movements such as chorea, dystonia, in-coordination and jerky eye movements.^19^ Despite the widespread reference to this hypothesis, we know of no direct neuroimaging evidence supporting it.

Establishing the role of altered basal ganglia connectivity in the pathogenesis of HD may also have a wider clinical relevance beyond simply understanding motor signs. Alongside the motor features of the condition, HD is associated with a marked psychiatric phenotype. Although associated with a range of psychiatric disturbances, there appears to be a unique relationship between HD and the development of apathy.^20^ Apathy, the loss of motivation and goal-directed behaviour, is highly prevalent in HD.^21^ Apathy in HD also tracks closely with disease progression even in premanifest and prodromal cohorts.^22^ Despite the high prevalence of apathy in HD however its pathogenesis is poorly understood and treatments are sorely lacking.^23,24^

Based on these epidemiological observations closely tying apathy to disease progression in HD, we hypothesised that like motor signs and apathy in early HD may also be a feature of basal ganglia pathway dysregulation. However, unlike motor signs, we hypothesised that apathy may instead arise from involvement of the *direct* basal ganglia pathway. We base this hypothesis on two strands of evidence. Firstly, as alluded to above, activation of direct pathway MSNs is thought to encourage free operant movement.^14–16^ The lack of free-operant action initiation is characteristic feature of behavioural apathy and disruption to the direct pathway may hamper this final stage of goal-directed behaviour – the expression of action.^25–27^ Secondly, computational models of basal ganglia function propose that as a result of the physiological asymmetry in dopamine receptor expression, these pathways not only play opponent roles in motor expression but also in goal-directed behaviour.^28–31^ Dysfunction in this pathway therefore may disrupt both the neural circuits necessary to take goal-direct action and impair the computational value associated with taking an action.

In summary, we sought to test two hypotheses – firstly that motor signs in HD would be associated with altered indirect pathway connectivity and secondly, our hypothesis that apathy in early HD may be associated with change in direct pathway connectivity.

To test these hypotheses, we used a neuroimaging technique to model direct and indirect pathway dysfunction. Canonically, these pathways are distinguished by change in activity that they *cause* within the thalamic nuclei. Within a neuroimaging framework, this causal connectivity is described as *effective connectivity*.^32^ Here we leverage the difference in both anatomical and effective connectivity to test our key hypotheses. To study effective connectivity, we use a Bayesian framework known as *dynamic causal modelling (DCM)* to build a simplified model of our pathways of interest.^33–36^ We based the model of these pathways on previous work in Parkinson’s disease (PD) but using several technological advances to test our hypotheses.^37^ Firstly, we used spectral DCM, a technique shown to outperform stochastic DCM for resting state fMRI data analysis.^38,39^ Secondly, at a group level, we used Parametric Empirical Bayes (PEB) to models how individual (within-subject) connections relate to between-subject factors such as motor scores.^40^ In this manner, our approach accounts for both expected values and model uncertainty at throughout our analysis. Finally, we did not specifically test our hypotheses but rather allowed an automated Bayesian procedure, called Bayesian Model Reduction (BMR). This approach allowed us to determine whether our hypothesised correlations between clinical scores and connectivity parameters emerged from the data *de novo*.^40–42^

Here we demonstrate, in a large cohort of patients with prodromal HD from the TRACK-ON HD study, that motor signs and apathy in HD are associated with unique basal ganglia connectivity profiles.^43^ Furthermore, we found that as hypothesised, higher motor scores were associated with connectivity changes in the indirect pathway components of our model. By comparison, higher apathy scores were associated with altered direct pathway connectivity changes.

## Material and Methods

### Sample

Data collected as part of the TRACK-ON HD were used in this analysis as previously described.^43^ For this analysis data from the third (and last) TRACK-ON visit were used. Participants aged below 18 or over 65 were not recruited and participants with major psychiatric, neurological, medical disorder or history of head injury were excluded. Participants with the HD mutation all had greater than or equal to 40 CAG repeats and a disease burden score of greater than 250 at baseline. The study was approved by the local ethics committees and all participants gave written informed consent according to the Declaration of Helsinki. Sample characteristics are described in Table 1. Of 102 scans which passed quality control, two participants were excluded for antipsychotic use. A further 6 participants who were left-handed were excluded leaving data from 94 HD gene carriers in the peri-manifest phase of the disease in this study. Although group differences were not the focus of this study, data from 85 right-handed control participants was also used to replicate baseline network connectivity as described below.

**Table 1:** sample demographics consisted of 94 HD gene carriers who underwent resting state fMRI as part of the TRACK-ON study. Apathy measured using the Baltimore Apathy Scale (BAS). Depressive symptoms measured using the Beck Depression Inventory (BDI). Spread of Unified Huntington’s Disease Rating Scale Total Motor Score (TMS) and apathy scores in supplementary figures. Table shows mean +/- std. unless otherwise stated.

|  |  |
| --- | --- |
| Age<br>(mean +/- std) | 45.5 (+/- 8.9) |
| % female | 50% |
| Mean CAG repeat<br>length | 43.1 (+/- 2.3) |
| TMS | 10.5 (+/- 8.5) |
| Apathy score | 10.9 (+/- 6.0) |
| Depressive scores | 6.6 (+/- 6.8) |
| Number by scanner<br>type (Siemens/Philips) | 52/42 |

### Clinical outcomes

Two primary outcomes were used in this study – Unified Huntington’s Disease Rating total motor score (TMS) and the self-rated Baltimore apathy scale (BAS).^44,45^ The motor score assesses the severity of 31 common neurological features such as chorea, dystonia, bradykinesia and oculomotor signs. The maximum score possible is 124. Due to the early stage of disease in these patients, and the relatively mild motor signs in the cohort (mean score 10.5 - see Table 1), the total motor score was used as opposed to specific subscales which would be underpowered. The BAS consists of 14 items with scores ranging from 0-42 where a higher score representing a higher degree of apathy. Self-rated apathy scores were used for this analysis. Self and carer rated apathy have good interrater reliability especially in the absence of significant cognitive impairment.^45–47^ To control for the effects of depression, Beck Depression Inventory scores (BDI) score were used as a covariate in the apathy analysis.^48^

### MRI data acquisition

3T MRI data were acquired at four sites: London, Paris, Leiden and Vancouver. T1-weighted image volumes were acquired using a 3D MPRAGE acquisition sequence as described by Kloppel *et al* (2015). For resting state fMRI, whole-brain volumes were acquired at a repetition time (TR) of 3s using a T_2_*-weighted echo planar imaging (EPI) sequence with the following parameters: echo time 30ms, field of view 212mm, flip angle 80°, 48 slices in ascending order (slice thickness: 2.8 mm, gap: 1.5 mm, in plane resolution 3.3 × 3 mm) and bandwidth of 1906 Hz/Px. In total 165 volumes were acquired over 8:20 min followed by field map acquisition.

### MRI pre-processing

MRI image pre-processing and quality control were as described in Kloppel *et al* (2015).^43^ In brief, the first four EPI images were discarded to allow for steady state equilibrium. Images were realigned and underwent inhomogeneity correction where field maps were available. EPI images were co-registered to anatomical images and normalised to MNI space. Data was smoothed with a 6mm full-width at half-maximum Gaussian kernel. Data underwent significant quality control as described by Kloppel *et al* (2015). Manual QC along with the use of ArtRepair and tsdiffana were used to assess for significant movement before pre-processing.

### Region of interest specification

A summary schematic of the pipeline used to analyse the rsfMRI data is shown in Fig 1. For this analysis, timeseries were extracted from four pre-defined regions of interest (ROIs) to make up the motor basal ganglia loop - the motor cortex, motor thalamus, motor putamen and the sub-thalamic nucleus (STN). With the exception of the subthalamic nucleus, timeseries were extracted from spheres seeded within anatomical masks defined in a standard space. An anatomical mask of Brodmann’s Area 4 from Wake Forest University Atlas was used to define the motor cortex.^49^ The motor putamen and motor thalamus masks were defined from probabilistic connectivity atlases with a threshold of 50% probability.^50,51^ The STN was not manually defined in this study. Instead, a mask made available by Keuken *et al* (2013) was used.^52^ This mask, defined in MNI space, was derived from the accurate high-resolution delineation of STN using 7T imaging, also assessing the impact of age. Based on the sample characteristics in this study (mean age 45.5 +/- 8.9), the mask for middle-aged individuals was used again with a conservative threshold of >50% probability. Given the size of this structure and the spatial resolution of functional imaging, the timeseries extracted may also contain signal from adjacent structures.

**Figure 1:**
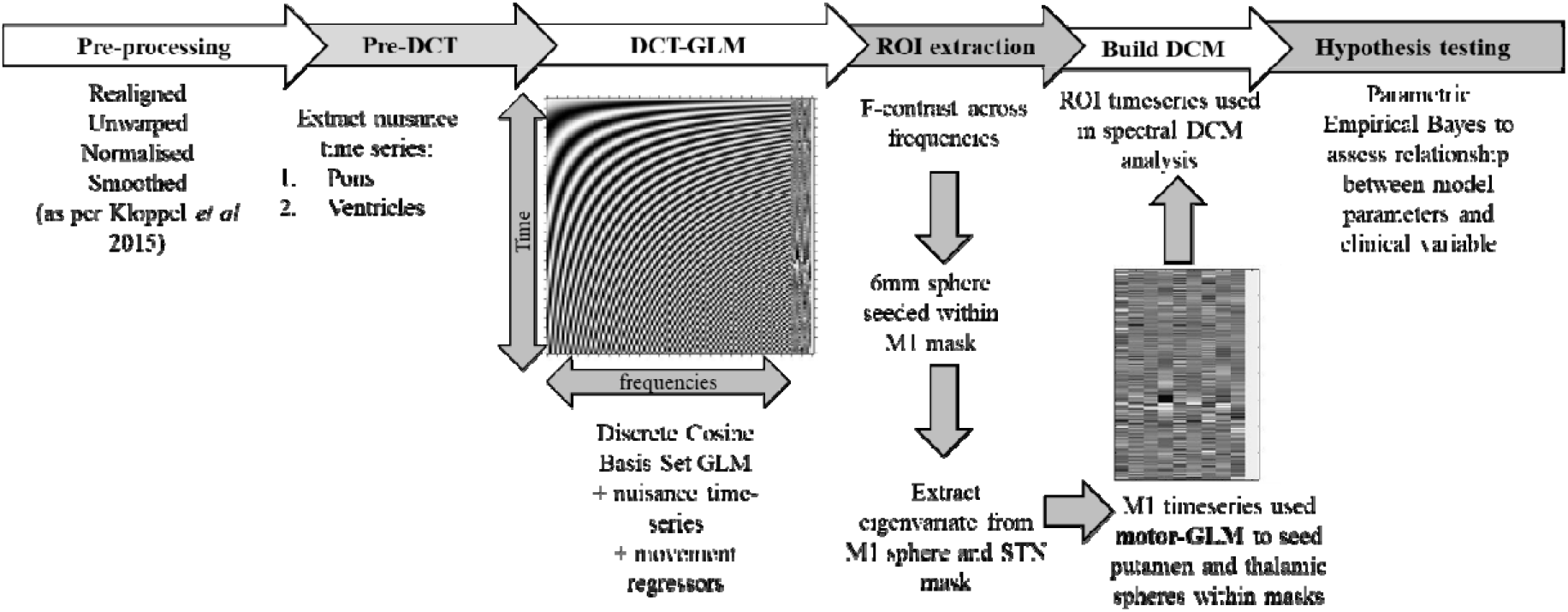
Summary of the rsfMRI analysis pipeline used in this study.

### Resting state fMRI modelling with GLM

Using the pre-processed scans, a dummy GLM was created to extract nuisance time series from the pons and ventricles. To better model resting state low frequency fluctuations, we then used a discrete cosine transform (DCT). In summary, this approach consists of 189 cosine basis functions modelling frequencies in the typical resting state range of 0.0078-0.1Hz.^53–57^ We created a GLM containing these DCT regressors as well as the nuisance timeseries extracted as described above alongside six movement regressors.

An F-contrast was used over the DCT frequencies to identify regions that showed resting state activity within the motor cortex. Based on this contrast, within the BA4 mask, a 6mm sphere was placed at the location which showed the highest activity in the frequencies of interest. From this sphere the principal eigenvariate (adjusting for head movements and nuisance timeseries) was extracted. This procedure summarises the timeseries from all of the voxels in the sphere into one representative timeseries for the ROI. The variance explained by the eigenvariate in the M1 ROI had a mean of 67% with a variance of +/- 11.5%. The principle eigenvariate from the entire sub-thalamic nucleus mask was also extracted as above with variance explained mean of 87% with a variance of +/- 4.3%. The timeseries extracted from the motor cortex was then used to determine the location of 4mm sphere placed within the motor putamen and motor thalamic masks. The centre of these spheres was placed within each mask, at the co-ordinates that showed the strongest correlation with the M1 timeseries regressor. The principal eigenvariate was extracted from these spheres controlling for the same confounders showing variance explained with a mean of 76% (variance: +/-8.6) and % (variance: +/-8.4) in the putamen and thalamus respectively. Example timeseries extracted from these ROIs is shown in Fig. S1.

### Dynamic casual modelling and specification of the connectivity matrix

Based on previously published work, we used a simplified circuit representing the direct, indirect and hyper-direct pathway as shown in Fig. 2.^54^ Here we do not model connections involving the globus pallidus, instead we use simplified circuit involving motor cortex, putamen, thalamus and STN as described by Kahan *et al* (2014). A forward connection fromM1 to motor putamen represents the input to the network from the motor cortex. Motor putamen was modelled as having two forward connections – one connecting it to the motor thalamus, forming the ‘direct pathway’ of our model, and a second connection linking it to the STN, the first component of the ‘indirect pathway’ of our model. The STN was modelled as having a further connection to the thalamus, forming the second connection within the model’s indirect pathway. A further direct connection between the cortex and the STN was specified representing the hyper-direct pathway. These basal ganglia pathways are shown as a schematic in Fig. 3A-C.

**Figure 2:**
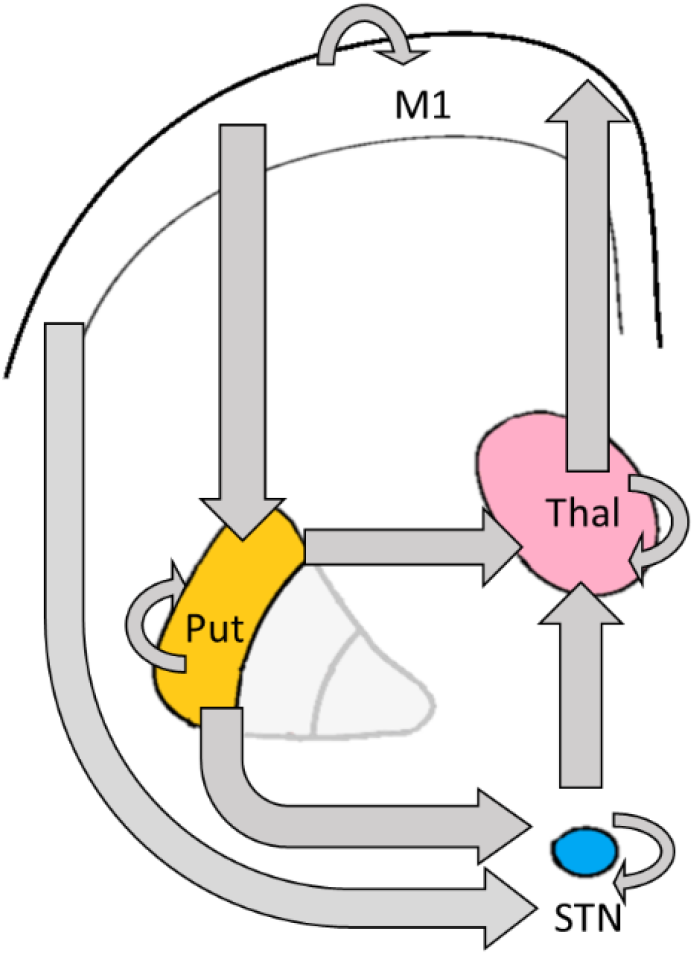
Schematic of the basal ganglia network modelled in this study (the DCM ‘A-matrix’). The direction of the arrows indicates that direction of effective connectivity entered into the model. Arrows looping back to the same node represent inhibitory self-connections specified in the DCM.

**Figure 3:**
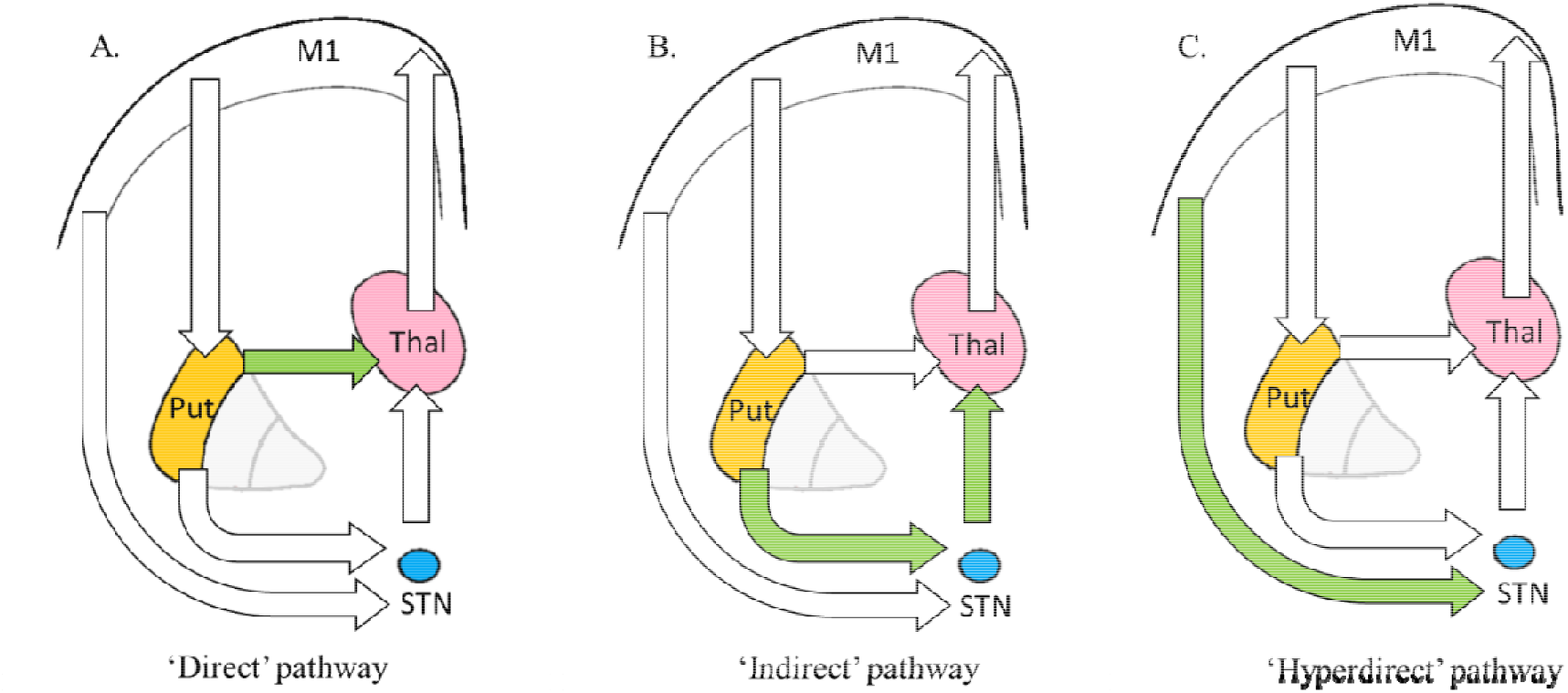
This model generates simplified representations of three pathways of interest. (A) The direct pathway is composed of the connection between putamen and thalamus. (B) The indirect pathway components are the putamen-STN connection and the STN-thalamic connection. (C) the hyperdirect pathway (C) composes of a connection from the motor cortex to the STN.

Having specified this network, or *A-matrix*, we used spectral DCM packaged as part of SPM12 to infer effective connectivity parameters. Unlike stochastic DCM, spectral DCM inversion does not predicts the time series extracted from each node but rather estimates their cross spectral density.^38^ This approach is more accurate at recovering parameters and estimated second-level effects such as group differences.^39^ In DCM, we do not specify the valence of the connections between nodes and allowed these to be estimated from the data. The positive connectivity value refers to an excitatory connection whereas negative connectivity value refers to an inhibitory influence.

### Hypothesis testing with parametric empirical Bayes (PEB) and Bayesian model reduction

The DCM specified above was estimated for each participant separately. The DCMs performed well with variance explained of 83.1% (variance: +/- 9.7%) in the patient cohort (and 83.8% (variance: +/-8.4%) in the control cohort). Inference on clinical scores was performed using Parametric Empirical Bayes (PEB).^40^ This is a between-participants hierarchical Bayesian model that models how connections at the individual level, such as connectivity parameters, relate to between subject factors, such as motor score. At the first level, individual participant parameters were estimated using spectral DCM. The PEB approach then considered these parameters at the second level as having group means and between participant variability which could be explained by between participant factors.

In this procedure, firstly a parent model – which can be sparse (as here) or fully connected - is estimated in which all regressors of interest such as motor score and covariates are modelled as having an effect on any of the connections specified in the subject level DCMs. In order to test our hypotheses, we combined this approach with *Bayesian Model Reduction* (BMR) procedure.^41,42^ BMR procedure scores all nested (reduced) models by turning off parameters that do not contribute to the *model evidence*. In brief, BMR enables the (greedy) search of very large model space by scoring each (reduced) model based on model evidence or free energy. The parameters of the best 256 reduced models from this search procedure are then averaged, weighted by their model evidence (i.e., Bayesian Model Averaging). For further details see ^41,42^. This procedure derives the posterior densities of the parameters by marginalising over the models accounting for model uncertainty. In this manner, parameter estimates are not heavily influenced by models with high levels of uncertainty.

Estimates of parameter strength are outputted along with the posterior probability of the parameters being non-zero. These parameters represent the rate of change in activity in the afferent node, measured in Hz, caused by activity in the efferent node. As described by Kahan *et al* (2014) they can be thought of as the sensitivity of the target node to the source. The PEB models we specified controlled for age, gender and scanner type. The effect of motor score and depressive scores were then additionally controlled for in analyses of apathy in the HD sample. Group comparison was not the focus of this study however data from control participants was used to replicate the baseline connectivity profile (as shown in Supplementary Material). Regressors were mean centred allowing the interpretation of the first covariate of the model to be the average connectivity weights in the network. We only report connections which have a posterior probability of >0.99 (which refers to very strong statistical evidence).

Having estimated the effect of clinical co-variates on connection strengths at a group level, we completed a Bayesian leave-one-out cross validation procedure, as implemented in SPM, to determine whether these weights could themselves be predictive of an individual participant’s symptom scores. Cross-validation of this sort provides out of sample estimates of predictability (i.e., the predictive validity of the connectivity strength from a new participant).^42^

## Results

### Sample demographics

Our sample consisted of HD gene carriers and controls recruited into the TRACK-ON study who had both clinical and neuroimaging data available. This cohort is peri-manifest with 34 of 94 patients having been diagnosed with early HD. In the early-stage HD cohort, the mean Total Motor Score (TMS) was 17.9 (+/- 9.2).

### Average connectivity parameters show a network supressing motor cortex activity

During data collection for resting state analysis, participants were explicitly asked to stay still across the scanning session. In keeping with this, average connectivity parameters showed active suppression of the driving input from the motor cortex. The net output from this system via the thalamocortical connection was to supress motor cortical activity (-0.39 Hz, 95% CI: -0.47 to -0.31 Hz, posterior probability (*pp*) > 0.99). The ‘direct pathway’ component of our model, the striato-thalamic connection, was found to be excitatory (0.43 Hz, 95% confidence interval (95% CI) 0.37 to 0.50 Hz, *pp* > 0.99). By comparison the two components of the ‘indirect pathway’ were found to be inhibitory: subthalamic-thalamic (-0.1 Hz, 95% CI: -0.15 to -0.04 Hz, *pp* > 0.99) and striato-subthalamic (-0.17 Hz, 95% CI: -0.24 to -0.11 Hz, *pp* > 0.99). These data are shown in a schematic in Fig. 4 with green arrows representing excitation, red arrows representing inhibition and grey arrows representing non-significant *effective* connectivity. This connectivity profile was also largely replicated in a cohort of control participants (*n* = 85) from the same study as shown in Figure S3 and Table S1.

**Figure 4:**
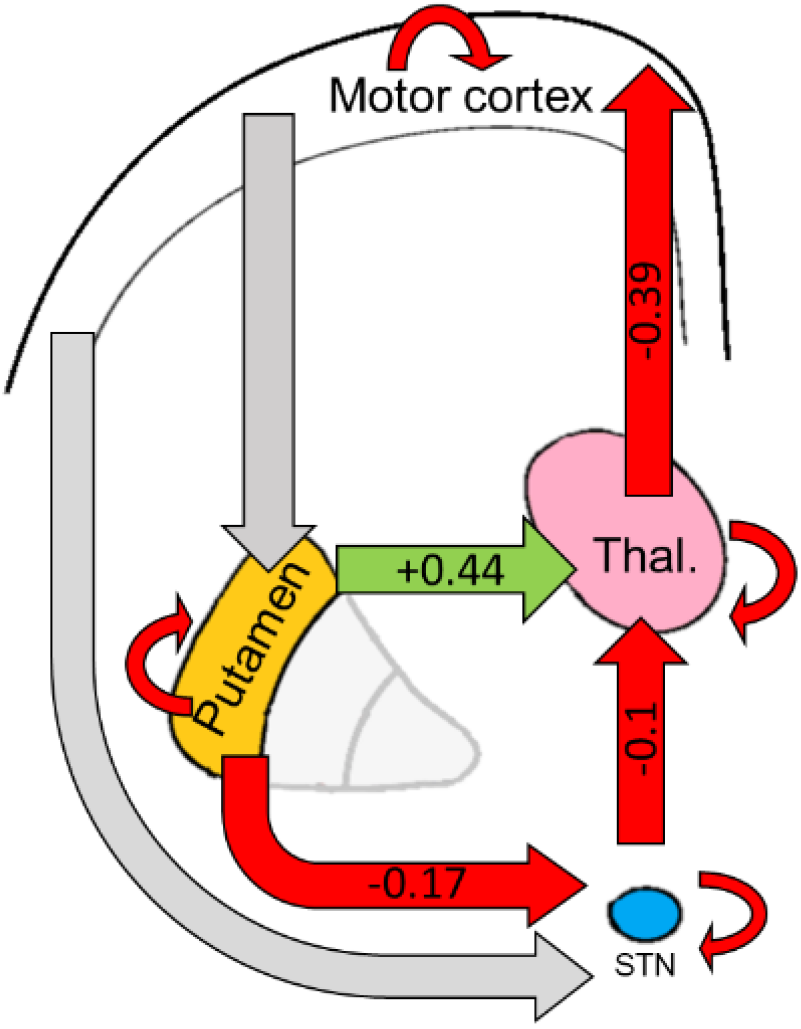
Schematic showing the average parameter values in the modelled network, across all 94 HD subjects, for between node connections. Red arrows indicate suppression of activity, green arrows indicate excitation and grey arrows indicate non-significant connections. Coloured arrows represent connections with a posterior probability of >0.99 for being greater than 0. Overall, the network activity shows a suppression of M1 activity which may be expected given that subjects are explicitly trying to remain still. Negative self-connections are shown as curved arrows looping back to the node – their values are described in Table S1. Model adapted from Kahan *et al* (2014).

### Altered connectivity basal ganglia connectivity associated with total motor scores and apathy scores

Using the PEB and BMR procedure (see Methods), we tested the hypothesis that changes in connectivity strength within our basal ganglia network would be associated with in total motor scores (TMS) and apathy scores. We report only connections found to have very strong evidence (posterior probability (pp) >0.99) of being associated with clinical scores. We went on to test whether the strength of these identified connections could predict clinical scores using a leave-one-out cross validation analysis. The results for motor and apathy analyses are shown graphically in Fig. 5A & 5B respectively. Both analyses control for age, sex and scanner type while apathy analysis also controls for depression and motor score. Results are given as normalised beta values with no units.

**Figure 5:**
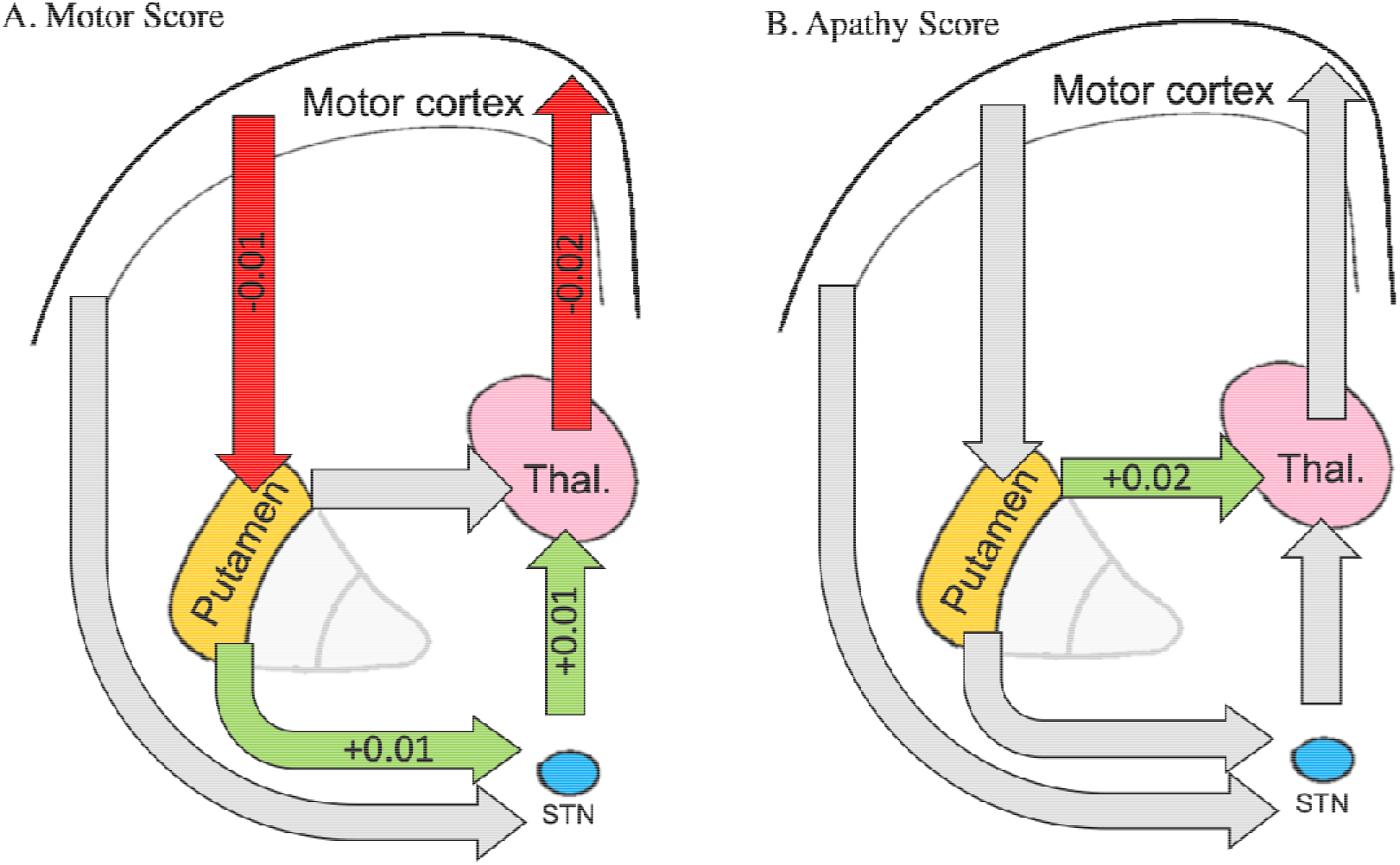
Association between inter-node connectivity parameters and (A) Total Motor Score and (B) Baltimore Apathy Score. Green and red arrows indicate which connections were found to be associated with clinical variable with >99% posterior probability using Parametric Empirical Bayes (PEB). Grey arrows show connections from connectivity matrix not found to be associated with clinical scores. Green arrows represent evidence of a positive relationship between connection strength and clinical scores, whereas are red arrows represent a negative relationship between clinical score and connection strength.

#### a. Motor score associated with changes in indirect pathway connectivity parameters

As shown in Fig. 5A, we found that total motor score (TMS) was positively associated change in both indirect pathway components of our model: striato-STN (0.013, 95% CI: 0.005 to 0.021, pp > 0.99) and STN-thalamic (0.011, 95% CI: 0. 005 to 0.018, pp > 0.99). With reference to Fig. 4, this means that as motor score increased, these connections became less inhibitory.

The weights from the two components of the indirect pathway of our model significantly predicted TMS (r = 0.17, p = 0.047) in a leave one-out-cross validation analysis.

TMS was also negatively associated with cortico-striatal connectivity (-0.009, 95% CI: - 0.014 to -0.004, pp > 0.99) and thalamo-cortical connectivity (-0.019, 95% CI: -0.030 to - 0.009, pp > 0.99). TMS was positively associated with STN self-connection (0.01, 95% CI: 0.006 to 0.016, pp > 0.99).

#### b. Apathy scores associated change in direct pathway connectivity scores

By comparison, total apathy score was positively associated with strength of the direct pathway component of our model, the striato-thalamic connection (0.022, 95% CI 0.014 to 0.03, pp > 0.99). With reference to Fig.4, this means that as apathy scores increased, the striato-thalamic connection becomes more excitatory.

Although strong evidence for this effect exists at a group level, weights of the striato-thalamic connection were not strong enough to predict individual apathy scores in a leave-one-out cross validation analysis (p = 0.30)

Apathy was also negatively associated with STN self-inhibition (-0.013, 95% CI: -0.018 to - 0.007, pp > 0.99).

## Discussion

We show using functional neuroimaging, that motor signs and apathy in HD are associated with unique profiles of altered *effective* connectivity within basal ganglia pathways. We found strong evidence at a group level that higher motor scores in a large cohort of peri-manifest HD patients were associated with altered coupling in the indirect pathway of our model. By comparison, we identified that apathy scores in prodromal HD may be associated with changes only in striato-thalamic or direct pathway connectivity within our model.

We found that motor signs were associated with less inhibition in the striato-STN and STN-thalamic components of our model whereas apathy was associated with increased coupling between putamen and thalamus. Although our hypotheses are based on rate-coding models of striatal function, given the limitation of interpreting BOLD signals we do not interpret our results as demonstrating of more or less activity in the cell populations we hypothesised. Instead, we simply report evidence that motor signs and apathy were associated with unique basal ganglia connectivity profiles with changes in connectivity associated with each clinical feature mapping onto the connections we hypothesised, within the confines of our model. Our findings may represent a range of pathological processes such as altered rating coding, synaptic dysfunction or altered basal ganglia synchrony.

Although the hypothesis that indirect pathway dysfunction drives the development of motor features of HD is well established, we know of no previous research neuroimaging research demonstrating a link between motor score and basal ganglia connectivity. Furthermore, in this paper we find evidence for novel hypothesis: that apathy in HD may also in part be driven by impaired basal ganglia connectivity, perhaps in the direct pathway. Activity in this pathway drives free-operant movement – a feature commonly lacking in apathy.^14–16^ Computational models of basal ganglia function argue that, via dopaminergic learning signals, the direct pathway cells effectively accrue the value of taking an action.^29–31^ Impaired coupling within this pathway may therefore disrupt both the striatal machinery necessary to take goal-directed actions and the neural representations of the value of those actions. Here we present evidence to support this novel hypothesis.

We believe this study has a number of design strengths. To test our hypotheses, we used data from a large cohort of HD gene carriers who were expressly recruited around the time of motor onset. Many motor signs in HD are not actively elicited and occur at rest – as such, resting state data has considerable ecological validity in trying to understand these features of the disease. The same may be said of apathy. In order to analyse this data, we used *spectral* DCM which has been shown to have several benefits when analysing resting state data.^38,39^ Using this technique, we found a network whose net output was to reduce activity in the motor cortex. There is little existing data with which to compare these results, but this profile was also replicated in a supplementary control cohort in a separate analysis. We tested the relationship between clinical variables and connections within the network using an advanced, Bayesian, approach.^40–42^ This procedure compares many competing hypotheses and only those with the strongest evidence survive. As such, our a priori predictions were not directly tested but were confirmed *de novo* from the data itself. In both analyses, we found very strong evidence to support our main hypotheses at a group level. In subsequent analyses we asked whether individual clinical scores could be predicted by the weights of the connections we identified at the group level. Using a leave-one-out cross validation procedure we found that only motor scores could be predicted from the connections strengths, not apathy scores.

We would also like to draw attention to a few limitations of this study. Firstly, in both analyses we found modest effect sizes. This is perhaps unsurprising. Firstly, in both cases we are sampling from a small region of each structure, and it is unlikely that all clinical change can be attributed to such a restricted region of interest. Secondly, many neural changes are associated with HD and the pathogenesis of both motor signs and apathy are likely to be biologically heterogenous. In the case of apathy in particular, multiple neurological mechanisms may contribute to the development of apathy in HD such as white matter changes, involvement of cortical structures or indeed the involvement of other striatal compartments, such as striasomes, which we are unable to currently resolve with *in vivo* imaging.^8,58–61^ We therefore do not claim, based on the data presented here that changes in connectivity that we present are sufficient to generate clinical features. Rather, we argue that changes in basal ganglia connectivity may contribute to their development in patients.

We should also highlight that we adopted a cross-sectional design. A longitudinal study would give a clearer understanding of the changes that drive the emergence of these features however this approach has a number of challenges. Given the slow rate at which clinical features evolve in HD, it is unlikely that longitudinal analysis over a few years would have sufficient power to detect changes in our areas of interest. Instead, we compared across participants with variance in relevant clinical features. We would hypothesise that similar results would be obtained longitudinally if sampled over a longer time period. Our cohort was also in the very earliest stages of manifest disease with low symptoms scores. Although this limited the variability in clinical scores, this cohort offered two key advantages. Firstly, very few participants needed to be excluded due to anti-dopaminergic medication use and secondly, participants at this stage of disease were able to tolerate fMRI imaging.

It is also clear that the model used this this study is a simplified model of the relevant basal ganglia circuits. Modelling the true extent of the anatomical complexity within basal ganglia circuits is currently intractable with fMRI and therefore any attempt to do so requires simplification.^62^ At the core of our model, also used by Kahan *et al* (2014), is a connection through which striatal activity can drive thalamic activity directly or via a secondary, indirect, route which necessitates striato-diencephalic connectivity in order to change thalamic connectivity.^54^ We found that these pathways excited and inhibited thalamic activity respectively. On this basis we described them as the *direct* and *indirect* pathways in our model however, we cannot confirm they represent activity in the MSNs as we hypothesise. Although a simplification, we believe this model sufficiently captures the principal dynamics of the network as relevant to the hypotheses we are testing, whilst also limiting model complexity. Finally, due to the size of the regions we were interested in, especially the subthalamic nucleus, partial volume effects are impossible to avoid. However, data extracted from these regions largely conformed to the pattern of activity expected for this system at rest namely, reduction of motor cortical activity, striato-thalamic excitation via the direct pathway and thalamic inhibition via the indirect pathway. In comparison, previous work treated the STN as a hidden node, meaning that activity from the region is simulated by the model based on *a priori* expected connectivity.^37,63^ Whilst avoiding partial volume effects, this approach has the limitation that the model itself must infer the timeseries from a key node in the network as opposed to modelling data taken from the region itself, the approach taken in this study.

In summary we demonstrate, using neuroimaging, that changes in connectivity in the basal ganglia motor loop are associated with motor sign severity and apathy in HD. In part the motivation for this study was to better inform the pathogenesis of these clinical features to advance therapeutics. For apathy in particular, our findings may suggest that medications which manipulate the relative activity of basal ganglia pathways, in particular those that modulate direct pathway activity, may be a fruitful way forward.

## Data Availability

on reasonable request

## Acknowledgements and Funding

We are grateful to the Track-ON HD investigators. Track-On HD was funded by the CHDI foundation, a not-for-profit organisation dedicated to finding treatments for Huntington’s disease.

A.N. is supported by the Leonard Wolfson Experimental Neurology Centre (Award: 525369).

A.R. is funded by the Australian Research Council (Ref s: DE170100128 and DP200100757) and Australian National Health and Medical Research Council Investigator Grant (Ref: 1194910).”

S.G. is supported by a Wellcome Trust Collaborative Award (200181/Z/15/Z).

R.B.R. is supported by a Medical Research Council Career Development Award (MR/N02401X/1) and a NARSAD Young Investigator Grant from the Brain & Behavior Research Foundation, P&S Fund. The Max Planck UCL Centre is a joint initiative supported by UCL and the Max Planck Society. The Wellcome Centre for Human Neuroimaging is supported by core funding from the Wellcome Trust (203147/Z/16/Z).

G.R. receives grant funding from the Wellcome Trust.

S.J.T. receives grant funding for her research from the Medical Research Council UK, the Wellcome Trust, the Rosetrees Trust, Takeda Pharmaceuticals, NIHR North Thames Local Clinical Research Network, UK Dementia Research Institute, Wolfson Foundation for Neurodegeneration and the CHDI Foundation.

We also recognise and would like to thank the participants for taking part in this study.

## Author Contributions

A.N. designed study, performed data analysis, wrote manuscript. Corresponding and Lead Author.

A.R. developed spectral DCM methods, contributed to the design of the study, helped significantly with data analysis, reviewed and amended manuscript.

S.G. helped with data analysis, reviewed and amended manuscript.

R.R. contributed to study design, provided valuable analysis guidance, reviewed manuscript.

G.R contributed to the design of the study, provided analysis guidance, reviewed manuscript.

S.J.T. contributed to the design of the study, provided analysis guidance, reviewed manuscript,

