## Supplementary File for "Imbalanced basal ganglia connectivity is associated with motor deficits and apathy in Huntington’s disease"

✉ denotes corresponding author

**(1)** Huntington's Disease Centre, UCL Queen Square Institute of Neurology, University College London **(2)** Max Planck UCL Centre for Computational Psychiatry and Ageing Research, UCL Queen Square Institute of Neurology, University College London. **(3)** Turner Institute for Brain and Mental Health, Monash Biomedical Imaging, Monash University **(4)** Department of Psychology, Yale University. **(5)** UCL Institute of Cognitive Neuroscience, University College London (6) Wellcome Centre for Human Neuroimaging, UCL Queen Square Institute of Neurology, University College London,

**Corresponding author:**

**(1)** Dr Akshay Nair, Huntington's Disease Centre, UCL Queen Square Institute of Neurology, University College London,

### Supplementary Material:

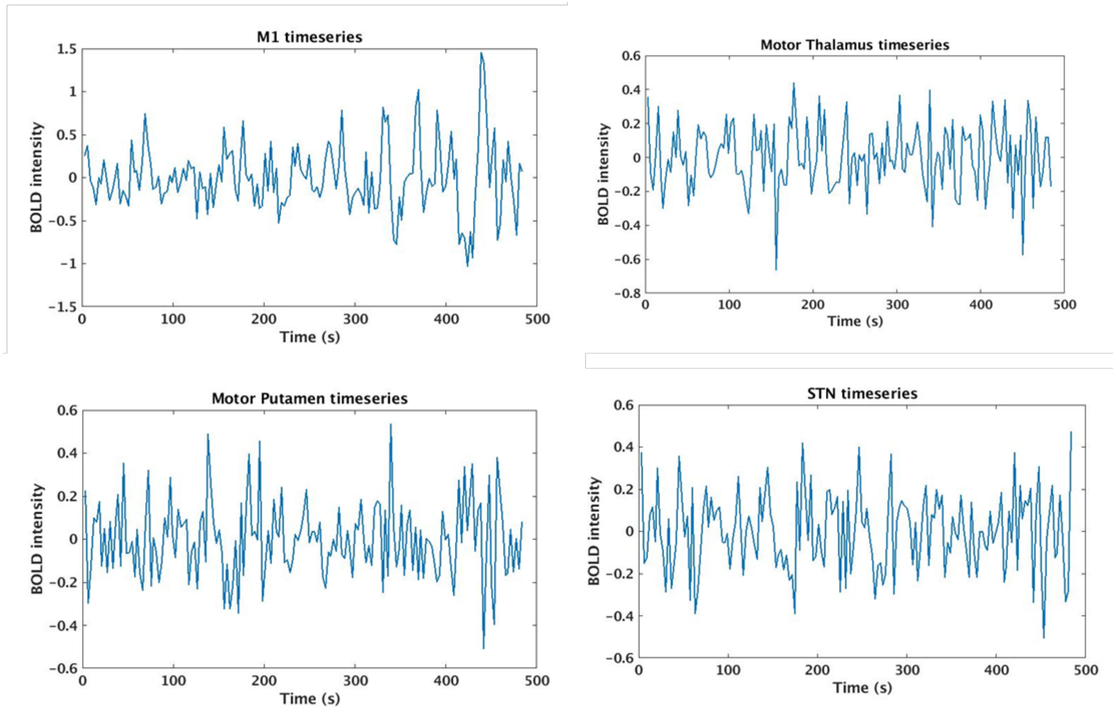

**Supplementary Fig (1):** Example timeseries from the four ROIs from one participant. Timeseries represent the principle eigenvariate of activity within voxels within each ROI. M1 timerseries is extracted from a 6mm sphere, putamen and thalamus timeseries are extracted from a 4mm sphere and subthalmic nucleus (STN) timeseries is extracted from an age appropriate mask.

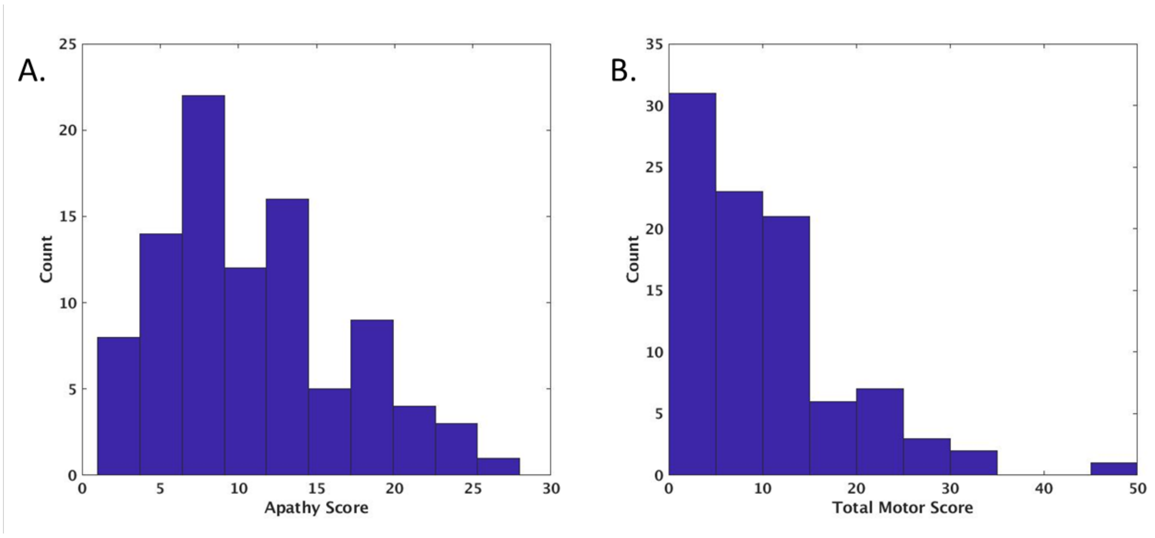

**Supplementary Fig (2):** A. Spread of apathy scores in the peri-manifest HD cohort as measured by the Baltimore Apathy Scale. Scores on this scale range from 0-42 with higher scores representing higher self-reported apathy. B. Spread of the Total Motor Score in the peri-manifest cohort as measured by the UHDRS total motor score. Scores on this scale range from 0-124. Relatively low scores in this cohort indicate early or premanifest disease.

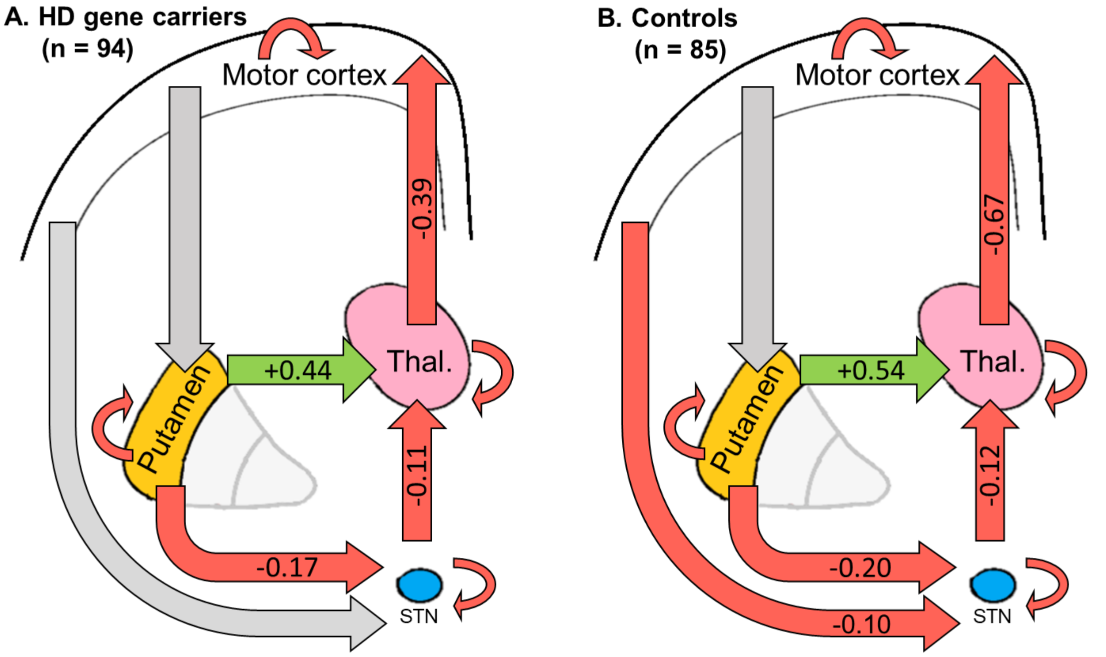

**Supplementary Fig (3):** Baseline connectivity estimates in the HD gene carriers showed active suppression of motor cortex at rest (model included TMS, age, gender and scanner type as covariates). This connectivity profile was replicated in a cohort of control participants (model included age, gender and scanner type). Parameter estimates with 95% CI and posterior probabilities are show in Supplementary Table 1 for both groups.

### Average coupling estimates show an active inhibition of M1 in HD gene carriers and controls

| | Gene carriers | | | | Controls | | | |
|---|---|---|---|---|---|---|---|---|
| **Connection** | **Mean value (Hz)** | **Lower 95% CI** | **Upper 95% CI** | **Post. prob** | **Mean value (Hz)** | **Lower 95% CI** | **Upper 95% CI** | **Post. prob** |
| **M1 to motor putamen** | 0.00 | 0.00 | 0.00 | 0.00 | -0.04 | -0.10 | 0.01 | 0.73 |
| **M1 to STN mask** | -0.05 | -0.12 | 0.01 | 0.79 | -0.10 | -0.15 | -0.05 | 1.00 |
| **Putamen to STN mask** | -0.17 | -0.24 | -0.11 | 1.00 | -0.20 | -0.26 | -0.14 | 1.00 |
| **Putamen to motor thalamus** | 0.44 | 0.37 | 0.50 | 1.00 | 0.54 | 0.47 | 0.62 | 1.00 |
| **STN to motor thalamus** | -0.10 | -0.15 | -0.05 | 1.00 | -0.12 | -0.17 | -0.07 | 1.00 |
| **Thalamus to M1** | -0.39 | -0.47 | -0.30 | 1.00 | -0.67 | -0.77 | -0.58 | 1.00 |
| **M1 self-connection** | 0.48 | 0.40 | 0.56 | 1.00 | 0.64 | 0.59 | 0.70 | 1.00 |
| **Putamen self-connection** | 1.16 | 1.11 | 1.22 | 1.00 | 1.08 | 1.02 | 1.15 | 1.00 |
| **STN self-connection** | 1.17 | 1.13 | 1.22 | 1.00 | 1.12 | 1.08 | 1.16 | 1.00 |
| **Thalamus self-connection** | 1.17 | 1.11 | 1.23 | 1.00 | 1.28 | 1.23 | 1.33 | 1.00 |

**Supplementary Table (1):** Comparison of baseline connectivity parameters in the motor network in gene carriers and controls. Both groups show the same overall pattern of effective connectivity (as shown in Supplementary Fig 5) which is suggestive that the motor cortex activity is being suppressed at rest. Positive values of self-connections reflect the degree of self-inhibition. Gene carrier values derive from a model including motor score, age, gender and scanner type. Control values derive from a model including age, gender and scanner type.
